# In COVID-19, *NLRP3* inflammasome genetic variants are associated with critical disease and these effects are partly mediated by the sickness symptom complex: a nomothetic network approach

**DOI:** 10.1101/2021.09.26.21264127

**Authors:** Michael Maes, Walton Luiz Del Tedesco Junior, Marcell Alysson Batisti Lozovoy, Mayara Tiemi Enokida Mori, Tiago Danellii, Elaine Regina Delicato de Almeida, Alexandre Mestre Tejo, Zuleica Naomi Tano, Edna Maria Vissoci Reiche, Andréa Name Colado Simão

## Abstract

**Background:** In COVID-19, the NLRP3 inflammasome is activated in response to SARS-CoV-2 infection. Acute infections are accompanied by a sickness symptom complex (SSC) which is a highly conserved symptom complex that protects against infections and hyperinflammation. Aims: To examine the associations of COVID-19, SSC and the *NLPR3 rs10157379 T>C* and *NLPR3 rs10754558 C>G* SNV variants; and the protective role of SSC in severe acute respiratory syndrome (SARS)-CoV-2 infection.

**Methods:** We recruited COVID-19 patients, 308 with critical, 63 with moderate and 157 with mild disease.

**Results:** Increased SSC protects against SARS, critical disease, and death due to COVID-19. Increasing age, male sex and *rs10754558 CG* significantly predict reduced SSC protection. The *rs10157379 CT* and *rs10754558 GG* genotypes are positively associated with SARS. Partial Least Squares analysis shows a) that 41.8% of the variance in critical COVID-19 symptoms could be explained by SSC and oxygen saturation (inversely associated), and inflammation, chest computed tomography abnormalities, increased body mass index, SARS and age (positively associated); and b) the effects of the *NLRP3 rs10157379* and *rs10754558* variants on critical COVID-19 are mediated via SSC (protective) and SARS (detrimental). SSC includes anosmia, dysgeusia and gastrointestinal symptoms.

**Conclusions:** Intersections among the *rs10754558 variant*, age, and sex increase risk towards critical COVID-19 by attenuating SSC. *NLRP3* variants play an important role in SARS, and severe and critical COVID-19 especially in individuals with reduced SSC, elderly people, and those with increased BMI, hypertension, and diabetes type 2. SSC is a new drug target to combat acute COVID-19.

## Introduction

Although the majority of COVID-19 patients have mild clinical manifestations, some patients may develop acute respiratory distress or even severe acute respiratory syndrome (SARS), which may necessitate intensive care unit (ICU) admission [1, 2]. SARS may progress into multiorgan failure and death particularly in the elderly and individuals with comorbid disorders, including type 2 diabetes mellitus (T2DM), hypertension, and obesity [2]. COVID-19 is caused by infection with the severe acute respiratory syndrome coronavirus 2 (SARS-CoV-2) and is accompanied by activation of immune-inflammatory pathways and sometimes with excessive inflammatory responses, which are associated with the severity of COVID-19 symptoms [2, 3]. Most importantly, during SARS-CoV-2 infection, the cytokine network is activated including increased levels of interleukin (IL)-1, IL-18 and tumor necrosis factor (TNF)-α in the peripheral blood [4, 5]. Severe COVID-19 is not only characterized by dysregulated release of these and other pro-inflammatory cytokines but also by acute lung injury and pneumonia, which can progress into SARS, disseminated intravascular coagulation, multisystem failure, and death [2]. In COVID-19, chest computed tomography scan anomalies (CCTA) and reduced peripheral capillary oxygen saturation (SpO2) are strongly associated with immune activation and inflammation [6]. It is thought that severe systemic inflammation with exuberant cytokine production, labeled as “cytokine storm,” is a primary cause of tissue injury in COVID-19, which may lead to SARS, organ failure and death [4, 5].

There is some evidence that SARS-CoV-2 stimulates the inflammasomes, a group of multimeric protein complexes which are responsive to pathogen-associated molecular patters (PAMPs), damage-associated molecular patterns (DAMPs) and environmental toxins [2, 7]. In COVID-19, the NLRP3 inflammasome is activated in response to SARS-CoV-2 infection as discovered in postmortem PBMCs and tissues of patients with moderate and critical COVID-19 [7,8,9]. NLRP3 or NOD, LRR (leucine rich repeat domain) and pyrin domain-containing protein 3 is an intracellular sensor which is primed by cellular stress via PAMPs (bacterial, viral and fungal infections) and DAMPs, resulting in the formation of the NLRP3 inflammasome with elevated levels of caspase-1, IL-1, and IL-18 and increased pyroptosis, an inflammatory form of cell death [9, 10].

NLRP3 is a key part of the innate immune system which orchestrates host-immune homeostasis and is involved in inflammatory and autoinflammatory disease [10]. Importantly, gain-of-function mutations in the NLRP3 gene may cause cryopyrin-associated periodic syndrome (CAPS), a dominantly inherited autoinflammatory disease characterized by recurrent episodes with systemic, musculoskeletal, cutaneous, and central nervous system inflammation [11, 12]. Moreover, the NLRP3 gene is associated with a large number of disorders and symptoms including viral disease, neurological disease, inflammatory bowel disease, multiple sclerosis, fever, myalgia, diabetes mellitus, Alzheimer’s disease, atherosclerosis, hypertension, heart disease, stroke, and obesity [12]. Nevertheless, the role of the *NLRP3* gene and the genetic variants including *NLPR3 rs10157379* and *NLPR3 rs10754558* in COVID-19 have remained elusive.

Acute infection is also accompanied by a sickness symptom complex (SSC) which is a symptom complex including fatigue, anergy, myalgia, hyperalgesia, malaise, listlessness, disinterest in social interactions, psychomotor retardation, anorexia, and weight loss [13, 14]. SSC is thought to be induced by acute infections and tissue injury through increased levels of IL-1 and TNF-α and is etiologically defined as an acute beneficial, adaptive response which plays an role in preventing the transition from the acute phase of inflammation to chronic inflammation by homeostatic processes including redirecting energy to activated immune cells and inducing a negative energy balance [13, 14]. As such, SSC is part of the compensatory immune-inflammatory system (CIRS), which tends to downregulate an overzealous acute inflammatory response [13, 14]. Chronic inflammation may develop following failure to eliminate the acute trigger, deficits in mounting SSC, persistent presence of innately chronic irritants, autoimmune responses to neoantigens, and activation of the Toll-Like Receptor (TLR) radical cycle [13–15]. The NLRP3/caspase 1 pathway plays a role in LPS-induced fatigue, suggesting that the NLRP3 inflammasome is involved in SSC [16]. Nevertheless, there are no data whether SSC may occur in response to SARS-CoV-2 infection or COVID-19 and whether a putative SSC response may protect against SARS, pneumonia, critical disease and death following SARS-CoV-2 infection.

Hence, the current study was performed to delineate a) the associations of COVID-19 outcome variables and SSC with the *NLPR3 rs10157379* and *NLPR3 rs10754558* genetic variants; and b) the protective role of SSC attenuating the more severe outcomes of SARS-CoV-2 infection including SARS symptoms, ICU admission and death due to COVID-19.

## Participants and Methods

### Participants

This cross-sectional study recruited 528 COVID-19 patients treated at the University Hospital of Londrina (HU) and the Emergency Rooms (ER) in Londrina, Paraná, Brazil. The WHO classification was employed to assess the clinical severity of COVID-19: 308 patients were classified as critical; 63 as moderate and 157 as mild [17]. Patients of both sexes who were over the age of eighteen were eligible. All ethnicities (self-reported) were included and registered as Caucasian or non-Caucasian. The presence of acute and chronic infections, cancer, and autoimmune diseases were exclusion criteria.

At the time of admission, a standard semi-structured interview was used to collect demographic, epidemiological, and anthropometric data from the COVID-19 patients, as well as clinical history, drug treatments, and comorbidities including type 2 diabetes mellitus (T2DM), hypertension, obesity, and hypothyroidism. We also registered intensive care admission (ICU), performance of orotracheal intubation, number of days from onset of illness to diagnosis, number of days from diagnosis until outcome of hospitalization, duration of acute illness, and death due to COVID-19.

At admission, we also registered the presence of a) SARS (yes/no) and SARS related symptoms, namely cough, dyspnoe, and fever; b) SSC symptoms including fatigue, muscle pain, loss of appetite, and headache; and c) diarrhea, gastro-intestinal symptoms, nausea, runny nose, dysgeusia, anosmia, and any neurological symptoms (all registered as yes/no). These data were used to compute severity scores of the SARS and SSC symptom profiles. SARS was conceptualized as the sum of SARS + dyspnoe + cough (labeled as SARSseverity) and SSC was conceptualized as the sum of fatigue + muscle pain + loss of appetite + headache (labeled as SSCseverity). Nevertheless, since we thought that also other symptoms may contribute to SARSseverity and SSCseverity we first examined the associations between those scales and other symptoms to delineate the final severity scores (see Results section). We also computed an index of severity of critical disease as the first principal component extracted from ICU admission, performance of orotracheal intubation, presence of SARS and death due to COVID-19 (labeled as DIIS index). All those items scored highly on the first factor (>0.654) which explained 61.65% of the variance (Kaiser-Meyer-Olkin measure of sampling adequacy=0.725). Body mass index (BMI) was calculated by dividing weight (kg) by height (m) squared.

The protocol was approved by the Institutional Research Ethics Committee of the University of Londrina, Paraná, Brazil (CAAE:31656420.0.0000.5231) and all of the participants and their giardians were informed in detail about the research and gave written informed consent.

## Methods

At admission, a venous blood collection (20 mL) was sampled using EDTA anticoagulant and clot activator in serum Vacutainer System tubes (Becton-Dickinson, New Jersey, U.S). Plasma and serum Eppendorfs and buffy coat were kept at −80°C until thawed for assays. Measurements of CCTA were obtained from all subjects at Hospital adimission. Noncontrast chest CT scans were performed on BRYGHTSPEED / GE 16 chanels (GE-Healthcare-America: Milwaukee, USA). The CT features comprized: ground-glass opacities, consolidations with halo sign, presence of nodules, pleural effusion and lymphadenophathy, and pulmonary involvement rated on an ordinal scale, namely 0-25%, 25-50%, 50-75% and more than 75%.

White blood cell counts (WBC) and leukocyte differentiation were determined by light scattering and fluorescence analysis (BC-6800 Mindray, Nanshan, China). Consequently, the neutrophil / lymphocyte ratio (NLR) was computed and used as an index of immune activation (Maes et al., 2021). Ferritin levels were determined using a chemiluminescent immunoassay (ALINITY I Abbott, Illinois, EUA) (BC-6800 Mindray, Nanshan, China). High sensitivity C-reactive protein (CRP) levels were assayed using turbidimetry (Architect C8000, Abbott Laboratory, Abbott Park, IL, USA). Based on the CRP concentrations and NLR, we computed a new index reflecting activation of immune-inflammatory pathways as z value of CRP (z CRP) + z NLR (labeled as inflammation index).

### Extraction of genomic DNA and genotyping of IL18

Following the manufacturer’s instructions, genomic DNA was extracted from the buffy coat of peripheral blood cells using a resin column procedure (Biopur, Biometrix Diagnostika, Curitiba, Brazil). The concentration of DNA was determined using a NanoDrop 2000cTM spectrophotometer (ThermoScientific, Waltman, MA, USA) at 260 nm, and the purity of DNA was determined by the 260/280 nm ratio. Real-time polymerase chain reaction (qPCR) with the TaqMan® (Thermo Fisher Scientific, Waltham, Massachusetts, EUA) method was used to identify SNV NLPR3 rs10157379 (T>C) and SNV NLPR3 rs10754558 (C>G) variants.

#### Statistics

To examine the relationships between categorical variables, contingency tables were analyzed using the Chi-square test or Fisher exact test (where appropriate). Associations between two variables were assessed using Pearson’s product moment or point-biserial correlation coefficients. Using analysis of variance, we examined the differences in scale variables between groups (ANOVA). Toward this end, the patient group was divided into three different classes using a visual binning method based on the SSCseverity scores (see Results section). Protected Least Significant Difference (LSD) tests were used to assess multiple pair-wise differences. When needed, continuous data were transformed using logarithmic (Ln) or square root transformations to normalize the distribution or to account for variance heterogeneity between study groups (as assessed with the Levene test). The primary outcome analyses were ANOVAs with SSCseverity and SARSseverity as dependent variables and the genotypes as explanatory variables. These multiple associations were subjected to p-correction for false discovery rate (FDR) [18]. When significant, we also examined the associations between the SSC and SARS and the genotypes using automatic stepwise binary logistic regression analyses. Consequently, we examined the effects of the genetic variants on these binary outcome variables considering other significant predictors (age, sex, comorbidities, CCTA, SpO2, and inflammation index). Toward this end we used automatic stepwise binary and multiple regression analyses. The results of these multivariable binary regressions were expressed as adjusted odds ratio (OR) with a 95% confidence interval (CI) and Nagelkerke values were used as pseudo R^2^ effect sizes. Automatic multiple regression analysis was used to predict dependent variables (SSCseverity, SARSseverity and DIIS scores) using biomarkers and demographic data (SpO2, CCTA, inflammation index, genetic variants, age, sex, BMI, etc) while checking for R^2^ changes, multivariate normality (Cook’s distance and leverage), multicollinearity (using tolerance and VIF), and homoscedasticity (using White and modified Breusch-Pagan tests for homoscedasticity). We used an automatic stepwise (step-up) method with p-to-enter of 0.05 and p-to-remove of 0.06. These regression analyses’ results were always bootstrapped using 5.000 bootstrap samples, and the latter results are shown if the results were not concordant. All tests are two-tailed, and statistical significance was determined using a p-value of 0.05. Factor analysis was performed, and the first extracted latent vector was relevant when all loadings were >0.666 and the variance explained > 50.0%. Factorability was checked using the Kaiser-Meier-Olkin (KMO) measure of sampling adequacy. IBM SPSS Windows version 25, 2017 was used for statistical analysis.

The multi-step multiple mediation associations between input variables (sex, genetic variants, age, CCTA, SpO2, SSCseverity, SARSseverity, inflammation index and BMI) and the output variables (severity of COVID-19 or critical disease) were assessed using Partial Least Squares-SEM analysis, namely PLSsmart [19]. All the input variables were entered as single indicators, while the output variables were entered as latent vectors (reflective models) extracted from several indicators. Complete PLS path analysis was performed using 5.000 bootstrap samples only when the outer and inner models met the following quality criteria: a) model quality SRMR index < 0.08; b) outer model loadings on the latent vectors > 0.666 at p < 0.001; c) the latent vectors show accurate construct validity as indicated by average variance extracted > 0.5, Cronbach’s alpha > 0.7, rho A > 0.8, and composite reliability > 0.8. To investigate compositional invariance, Predicted-Oriented Segmentation analysis, Multi-Group Analysis, and Measurement Invariance Assessment were used. PLSpredict with 10-fold cross-validation was used to assess the model’s predictive performance.

## Results

### Socio-demographic data

**Table 1** shows the socio-demographic, clinical and biomarker data of COVID-19 patients divided according to SSCseverity using a visual binning method which divided the sample into three groups (using > 7.2 and >9.3 as cutoff points) resulting in groups with minimal (minSSC), moderate (modSSC) and high (highSSC) SSC. We found significant point-biserial correlations between the sum of the 4 key SSC symptoms (fatigue, loss of appetite, myalgia, headache) and anosmia (r=0.397, p<0.001) and dysgeusia (r=0.375, p<0.001). Factor analysis showed that dysgeusia, anosmia and the four SSC symptoms loaded highly (all>0.679) on the first factor which explained 66.4% of the variance (KMO=0.621). Consequently, we computed the sum of six symptoms to reflect SSCseverity. The latter was strongly associated with the sum of the four key SSC symptoms (r=0.903, p<0.001, n=528). There were no differences in any of the statistical results shown below between SSCseverity computed based on those four or six symptoms. Interestingly, SSCseverity was also associated with diarrhea (r=0.299, p<0.001, n=528), nausea (r=0.177, p<0.001), and gastro-intestinal symptoms (r=0.321, p<0.001). Moreover, factor analysis showed that one relevant latent vector may be extracted from SSCseverity (0.815) and the sum of the three GIS symptoms (0.815) and that this factor explained 66.4% of the variance (KMO=0.609). The SSCseverity score was not associated with fever (r=-0.085, p=0.052) and was inversely associated with cough (r=-0.181, p<0.001), dyspnea (r=-0.286, p<0.001), SARS (r=-0.279, p<0.001), and SARSseverity (r=−0.368, p<0.001). Also, in patients with critical disease there was an inverse association between SSCseverity and SARSseverity (r=-0.181, p=0.001, n=308).

**Table 1.** Sociodemographic data, symptoms, and laboratory parameters of COVID patients divided into three groups according to severity of sickness symptom complex (SSC).

| Variables | COVID-19 + minSSC <sup>A</sup><br>n=241 | COVID-19 + modSSC <sup>B</sup><br>n=152 | COVID-19 + highSSC <sup>C</sup><br>n=135 | F/X<sup>2</sup> | df | p |
| --- | --- | --- | --- | --- | --- | --- |
| Sex (Female / Male) | 94/147 <sup>C</sup> | 73/79 | 78/57 <sup>A</sup> | 12.49 | 2 | 0.002 |
| Age (years) | 64.6 (16.8) <sup>B,C</sup> | 58.6 (1.5) <sup>A,C</sup> | 47.5 (17.3) <sup>A,B</sup> | 40.77 | 2/525 | <0.001 |
| Number days until diagnosis (days) | 6.1 (4.4) | 6.4 (3.4) | 7.4 (4.9) | 2.39 | 2/403 | 0.093 |
| Days from diagnosis until outcome of hospitalization (days) | 15.5 (13.0) | 14.4 (8.2) | 14.6 (8.5) | 0.45 | 2/401 | 0.641 |
| Duration of acute phase (days) | 21.6 (14.9) | 20.9 (9.8) | 22.0 (10.7) | 0.19 | 2/401 | 0.828 |
| Ethnicity (C/NC) | 195/46 | 122/29 | 104/31 | 0.92 | 2 | 0.632 |
| Smoking (N/Y) | 228/13 | 149/3 | 131/4 | 3.33 | 2 | 0.189 |
| Body mass index (kg/m<sup>2</sup>) | 27.72 (5.80) | 27.82 (5.28) | 27.90 (5.59) | 0.04 | 2/379 | 0.964 |
| T2DM (on oral hypoglycemics) (N/Y) | 173/68 <sup>C</sup> | 114/37 <sup>C</sup> | 118/17 <sup>A,B</sup> | 12.09 | 2 | 0.002 |
| Hypertension (N/Y) | 120/121 <sup>C</sup> | 75/77 <sup>C</sup> | 100/35 <sup>A,B</sup> | 24.38 | 2 | <0.001 |
| Fatigue (N/Y) | 198/42 <sup>B,C</sup> | 82/70 <sup>A,C</sup> | 37/98 <sup>A,B</sup> | 111.41 | 2 | <0.001 |
| Sickness symptom complex severity | 9.36 (0.48) <sup>B,C</sup> | 11.39 (0.49) <sup>A,C</sup> | 14.22 (1.25) <sup>A,B</sup> | 1973.2<br>3 | 2/525 | <0.001 |
| SARS (N/Y) | 138/103 <sup>B,C</sup> | 110/42 <sup>A,C</sup> | 114/21 <sup>A,B</sup> | 31.10 | 2 | <0.001 |
| SARS severity | 4.87 (0.88) <sup>B,C</sup> | 4.51 (0.98) <sup>A,C</sup> | 4.07 (0.89) <sup>A,B</sup> | 33.07 | 2/525 | 0.415 |
| DIIS index | 0.110 (1.032) <sup>B,C</sup> | -0.118 (0.955) <sup>A</sup> | -0.192 (0.917) <sup>A</sup> | 3.31 | 2/403 | 0.038 |
| Inflammation index | -0.011 (1.112) | 0.305 (1.015) | -0.020 (0.746) | 0.14 | 2/525 | 0.843 |
| C-Reactive Protein (CRP) mg/L * | 126.4 (90.7) | 130.9 (99.1) | 121.5 (81.5) | 0.03 | 2/525 | 0.975 |
| Neutrophil/Lymphocyte ratio | 10.32 (9.61) | 10.63 (9.97) | 10.63 (9.97) | 0.24 | 2/525 | 0.791 |
| Ferritin (ng/mL) * | 1533 (3075) | 1404 (1271) | 4035 (20693) | 2.37 | 3/391 | 0.095 |
| CCTA | 3.17 (1.20) | 3.34 (1.01) | 3.22 (0.71) | 0.88 | 2/521 | 0.415 |
| SpO2 (%) | 87.3 (7.1) | 87.9 (6.4) | 87.8 (3.7) | 0.59 | 2/525 | 0.557 |
| Mild / Moderate + critical COVID-19 | 35/206 <sup>B,C</sup> | 41/111 <sup>A,C</sup> | 81/54 <sup>A,B</sup> | 86.43 | 2 | <0.001 |
| Intensive care unit (N/Y) | 159/82 <sup>C</sup> | 112/40 | 113/22 <sup>A</sup> | 13.81 | 2 | 0.001 |
| Orotracheal intubation (N/Y) | 163/78 <sup>C</sup> | 119/33 <sup>C</sup> | 124/11 <sup>A,B</sup> | 28.80 | 2 | <0.001 |
| Death (N/Y) | 134/95 <sup>B,C</sup> | 89/31 <sup>A</sup> | 47/10 <sup>A</sup> | 16.24 | 2 | <0.001 |

All results of Chi-square tests ( $X^2$ ) or analysis of variance (F). FEPT: results of Fisher's exact probability test. Results are expressed as mean ( ± SD), <sup>A,B,C</sup>: multiple comparisons among treatment means, \* Processed in Ln transformation
C: Caucasian, NC: Not Caucasian, T2DM: type 2 Diabetes mellitus, DIIS index: first factor extracted from death, intensive care unit admission, orotracheal intubation, SARS: severe acute respiratory syndrome, CCTA Chest CT scan anomalies, SpO2: oxygen saturation percentage, Inflammation index: computed as a z unit weighted composite score of z CRP + z neutrophil/lymphocyte ratio

We found significantly more females in the highSSC as compared with the minSSC group. Age was significantly different between the three subgroups and decreased from minSSC to modSSC to highSSC. There were no significant differences in number of days until diagnosis, days from diagnosis until outcome of hospitalization and duration of the acute phase between the three subgroups. We found no significant associations between duration of illness and SSC or SARSseverity in the total study group and in the mild, moderate, and critical COVID-groups. There were no significant differences in ethnicity, smoking and BMI between the three groups. There were no significant differences in the prevalence of obesity (χ2=3.50, df=2, p=0.174), asthma (χ2=0.93, df=2, p=0.629), stroke (χ2=5.51, df=2, p=0.064), COPD (χ2=0.56, df=2, p=0.076), chronic kidney disease (χ2=3.02, df=2, p=0.221), hypothyroidism (χ2=5.92, df=2, p=0.052), and cardiac insufficiency (χ2=2.98, df=2, p=0.226) between the three study groups. Patients belonging to the highSSC group showed a lower incidence of T2DM and hypertension than the other two groups.

The prevalence of fatigue (as part of SSC) significantly increased from the minSSC to the modSSC to the highSSC group. The prevalence of SARS (no/yes) and SARSseverity were significantly different between the three groups and decreased from minSSC to modSSC to highSSC. The DIIS index was significantly lower in people with modSSC and highSSC than in the minSSC group. There were no associations between the immune-inflammatory markers (CRP, NLR, ferritin), CCTA and SpO2 and the three SSC groups. The mild / moderate + critical COVID-19 ratio was significantly different between the three SSC groups and increased from minSSC → modSSC → highSSC. Patients belonging to the highSSC group showed a lower incidence of ICU admission as compared with the minSSC group. Treatment with orotracheal intubation was significantly lower in the highSSC group than in the two other groups. Mortality was significantly lower in patients with ModSSC and highSSC as compared with MinSSC patients.

### Associations between inflammasome genetic fractions and SARSseverity and SSCseverity

Both genotypic distributions were in Hardy-Weinberg equilibrium, namely *NLPR3 rs10157379*: X2=2.61, df=1, p=0.106, and *NLPR3 rs10754558*: X2=1.91, df=1, p=0.167. **Table 2** shows the primary outcome analyses, namely ANOVA analyses performed on SARSseverity and SSCseverity with both SNVs as explanatory variables. As such, 4 ANOVAs were conducted, two were significant (shown in table 2) and two not. The significance levels presented in Table 2 remained significant after FDR p-correction (at p=0.036). There is a significant association between the *NLPR3 rs10157379* genetic variant and SARSseverity with significantly higher scores in the CT genotype as compared with CC and CC + TT combined. There was also a significant association between the *NLPR3 rs10754558* SNV and SSCseverity with lower scores in the CG genotype than in GG and CC.

**Table 2.**
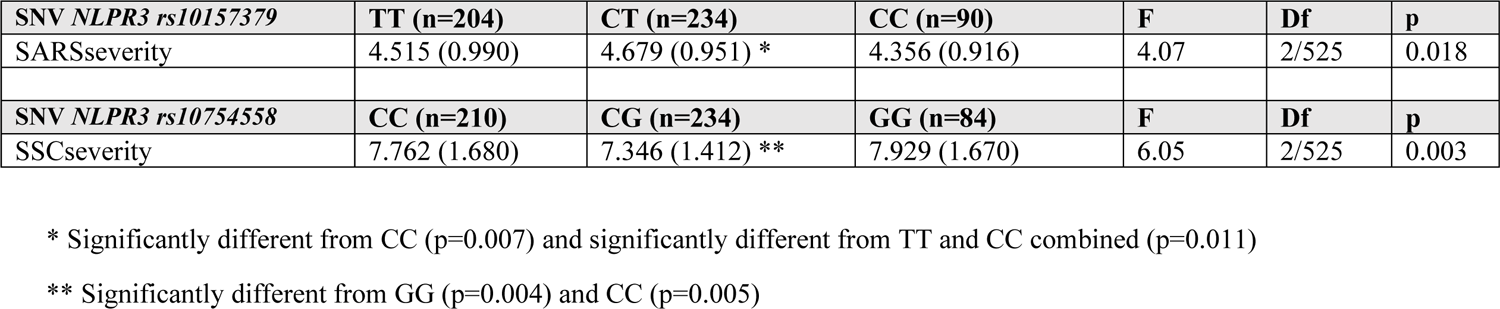
Results of univariate GLM analysis showing the differences in severity of the sickness symptom complex (SSCseverity) and severe acute respiratory syndrome (SARSseverity) between *NLRP3* inflammasome genotypes

| **SNV *NLRP3* rs10157379** | **TT (n=204)** | **CT (n=234)** | **CC (n=90)** | **F** | **Df** | **p** |
| --- | --- | --- | --- | --- | --- | --- |
| SARSseverity | 4.515 (0.990) | 4.679 (0.951) * | 4.356 (0.916) | 4.07 | 2/525 | 0.018 |
| **SNV *NLRP3* rs10754558** | **CC (n=210)** | **CG (n=234)** | **GG (n=84)** | **F** | **Df** | **p** |
| SSCseverity | 7.762 (1.680) | 7.346 (1.412) ** | 7.929 (1.670) | 6.05 | 2/525 | 0.003 |

\* Significantly different from CC (p=0.007) and significantly different from TT and CC combined (p=0.011)
\*\* Significantly different from GG (p=0.004) and CC (p=0.005)

Following these primary analyses, we also performed explorative analyses checking the associations between the separate symptoms and the genetic variants (namely the *NLPR3 rs10157379* genotypes and SARS symptoms and the *NLPR3 rs10754558* genotypes and the SSC symptoms). **Table 3** shows the results of logistic regression analyses and that SARS, cough, runny nose, and neurological symptoms were associated with the *NLPR3 rs10157379* SNVs. Binary multivariate logistic regression analysis showed that both the *NLPR3 rs10157379 CT* and *NLPR3 rs10754558 GG* genotypes predicted SARS symptoms. Fatigue, myalgia, loss of appetite, and headache were associated with the SNV *NLPR3 rs10754558*.

**Table 3.**
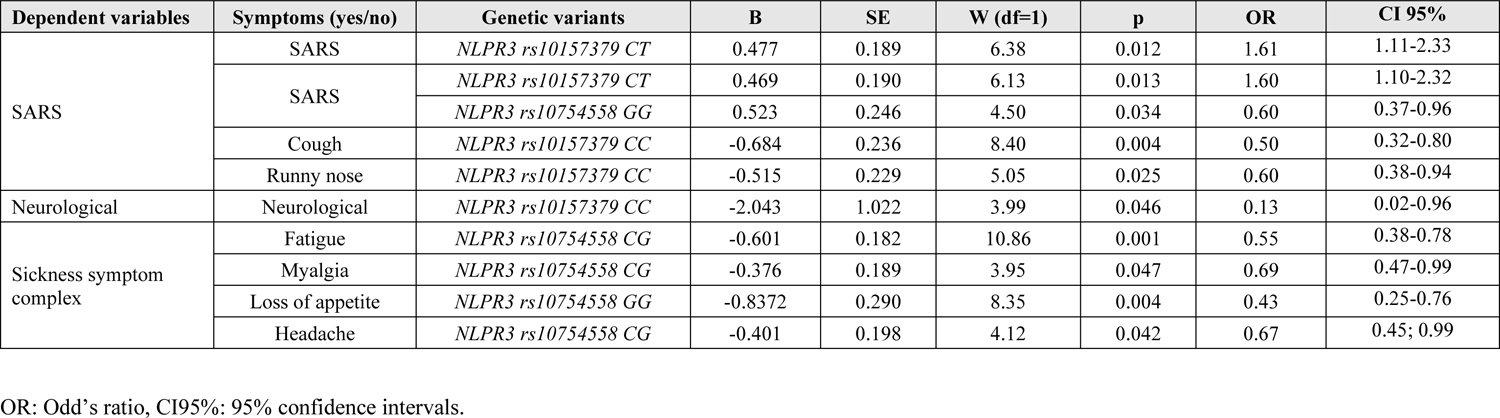
Associations between *NLRP3* inflammasome genetic variants and symptoms of severe acute respiratory syndrome (SARS) and sickness behavior.

| Dependent variables | Symptoms (yes/no) | Genetic variants | B | SE | W (df=1) | p | OR | CI 95% |
| --- | --- | --- | --- | --- | --- | --- | --- | --- |
| SARS | SARS | *NLRP3 rs10157379 CT* | 0.477 | 0.189 | 6.38 | 0.012 | 1.61 | 1.11-2.33 |
|  | SARS | *NLRP3 rs10157379 CT* | 0.469 | 0.190 | 6.13 | 0.013 | 1.60 | 1.10-2.32 |
|  |  | *NLRP3 rs10754558 GG* | 0.523 | 0.246 | 4.50 | 0.034 | 0.60 | 0.37-0.96 |
|  | Cough | *NLRP3 rs10157379 CC* | -0.684 | 0.236 | 8.40 | 0.004 | 0.50 | 0.32-0.80 |
|  | Runny nose | *NLRP3 rs10157379 CC* | -0.515 | 0.229 | 5.05 | 0.025 | 0.60 | 0.38-0.94 |
| Neurological | Neurological | *NLRP3 rs10157379 CC* | -2.043 | 1.022 | 3.99 | 0.046 | 0.13 | 0.02-0.96 |
| Sickness symptom complex | Fatigue | *NLRP3 rs10754558 CG* | -0.601 | 0.182 | 10.86 | 0.001 | 0.55 | 0.38-0.78 |
|  | Myalgia | *NLRP3 rs10754558 CG* | -0.376 | 0.189 | 3.95 | 0.047 | 0.69 | 0.47-0.99 |
|  | Loss of appetite | *NLRP3 rs10754558 GG* | -0.8372 | 0.290 | 8.35 | 0.004 | 0.43 | 0.25-0.76 |
|  | Headache | *NLRP3 rs10754558 CG* | -0.401 | 0.198 | 4.12 | 0.042 | 0.67 | 0.45; 0.99 |

OR: Odd's ratio, CI95%: 95% confidence intervals.

### Prediction of fatigue and SARS and outcome assessments

**Table 4** shows the outcome of different binary logistic regression analysis with fatigue and SARS as outcome variables and age, sex, ethnicity, smoking, BMI, comorbid diseases, genotypes, SpO2, and CCTA and the immune-inflammatory biomarkers as explanatory variables. We found that fatigue was significantly predicted by sex (higher in females), the *NLPR3 rs10754558 CG* genotype (protective), smoking and hypothyroidism (both inversely associated). SARS was significantly predicted by eight input variables with SSC being the most significant predictor (inversely associated). The other predictors were age, CCTA, CRP, *NLPR3 rs10754558 GG* and *NLPR3 rs10157379 CT* and T2DM (all positively associated) and SpO2 (inversely associated). Moderate and critical (versus mild) COVID-19 was significantly predicted by age, ferritin, SARSseveity, T2DM, and hypertension (all positively associated) and SSCseverity (inversely associated).

**Table 4.**
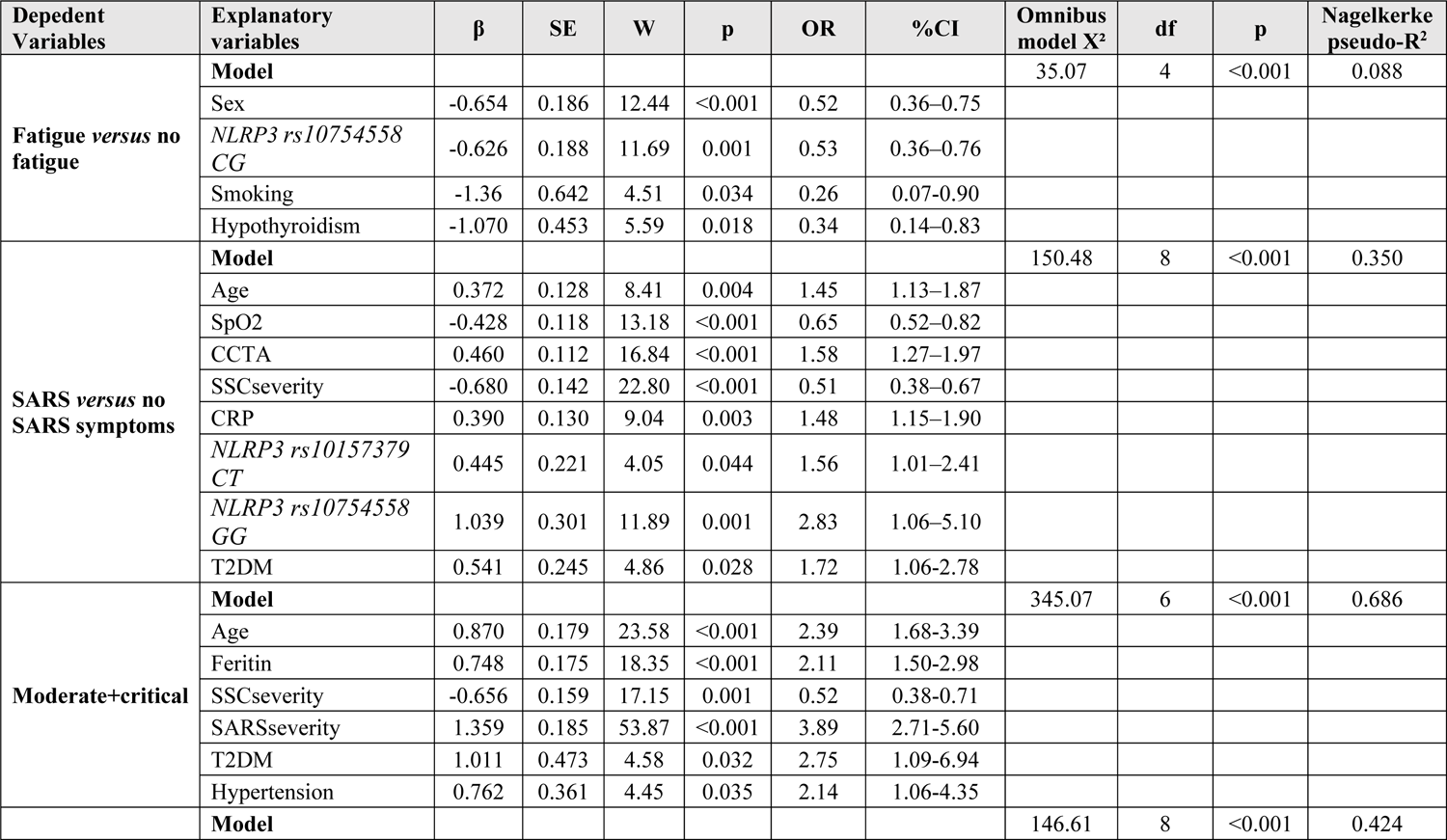
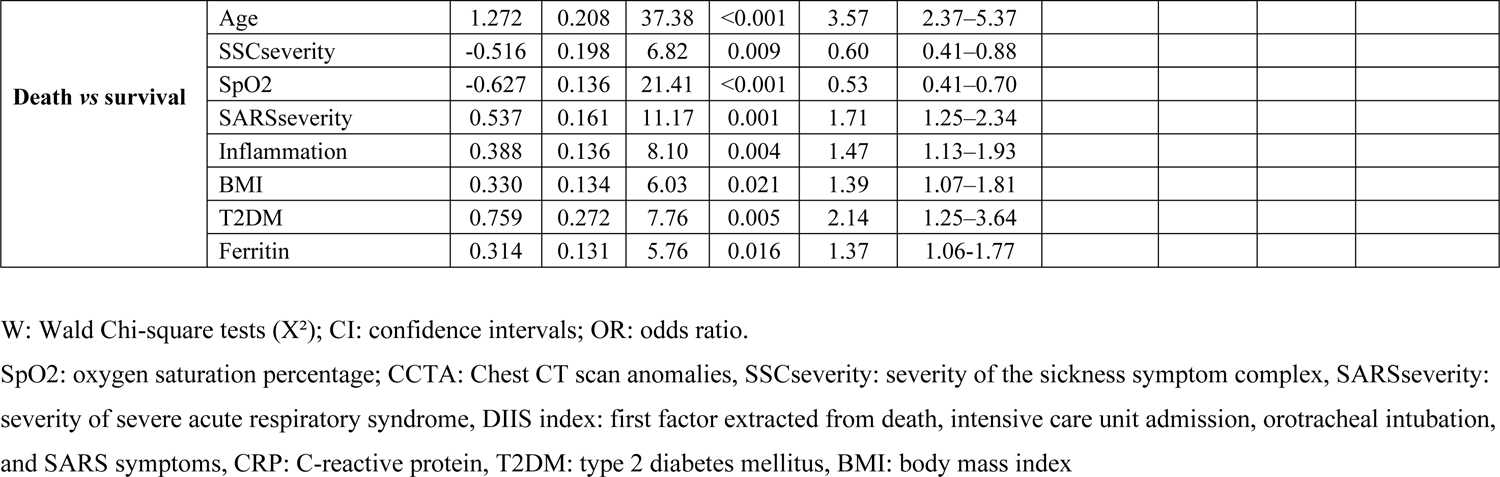
Results of binary logistic regression analysis with clinical outcomes as dependent variables

In the total study group, we found significant point-biserial correlations between death and SSCseverity (r=-0.184, p<0.001, n=406) and SARSseverity (r=0.254, n<0.001, n=406). Also, in the restricted study sample of critical COVID-19 patients, we found significant relationships between death and SSCseverity (r=-0.188, p<0.001, n=308) and SARSseverity (r=0.154, n<0.001, n=308). Table 4, shows that mortality was significantly predicted by age, SARSseverity, the inflammation index, BMI, T2DM and ferritin (all positively associated) and SSCseverity and SpO2 (both inversely associated).

### Prediction of SSCseverity, SARSseverity, and outcome assessments

**Table 5** displays the results of multiple regression analyses with SARSseverity, SSCseverity and DIIS index as dependent variables. We found that 25.5% of the variance in SARSseverity score could be explained by the regression on SSCseverity, SpO2, and *NLPR3 rs10157379 CC* genotype (inversely associated) and CCTA, age, and BMI (all positively associated). We found that 21.4% of the variance in SSCseverity was explained by age, and the *NLPR3 rs10754558 CG* genotype (both inversely associated) and female sex. Up to 24.5% of the variance in the DIIS index was explained by the regression on 6 input variables, namely SSCseverity, inflammation index, T2DM, CCTA and the *NLPR3 rs10157379 CT* genotype (all positively associated) and SpO2 (inversely associated).

**Table 5.** Results of multiple regression analyses with symptoms profiles and high-risk indices as dependent variables.

| Dependent Variables | Explanatory Variables | $\beta$ | t | P value | F | Df | p | R<sup>2</sup> |
| --- | --- | --- | --- | --- | --- | --- | --- | --- |
| **SARSseverity** | **Model** |  |  |  | 28.90 | 7/516 | <0.001 | 0.255 |
|  | SSCseverity | -0.303 | -7.23 | <0.001 |  |  |  |  |
|  | CCTA | 0.205 | 5.29 | <0.001 |  |  |  |  |
|  | Age | 0.152 | 3.59 | <0.001 |  |  |  |  |
|  | SpO2 | -0.144 | -3.71 | <0.001 |  |  |  |  |
|  | BMI | 0.111 | 2.90 | 0.004 |  |  |  |  |
|  | *NLRP3 rs10157379 CC* | -0.086 | -2.27 | 0.024 |  |  |  |  |
| **SSCseverity** | **Model** |  |  |  | 47.06 | 3/520 | <0.001 | 0.214 |
|  | Age | -0.409 | -10.49 | <0.001 |  |  |  |  |
|  | Female sex | 0.132 | 3.38 | <0.001 |  |  |  |  |
|  | *NLRP3 rs10754558 CG* | -0.127 | -3.27 | 0.001 |  |  |  |  |
| **DIIS index** | **Model** |  |  |  | 21.32 | 6/395 | <0.001 | 0.245 |
|  | SpO2 | -0.290 | -6.36 | <0.001 |  |  |  |  |
|  | Inflammation index | 0.176 | 3.79 | <0.001 |  |  |  |  |
|  | T2DM | 0.138 | 3.14 | 0.002 |  |  |  |  |
|  | CCTA | 0.150 | 3.26 | 0.001 |  |  |  |  |
|  | SSCseverity | -0.128 | -2.91 | 0.004 |  |  |  |  |
|  | *NLRP3 rs10157379 CT* | 0.096 | 2.00 | 0.046 |  |  |  |  |

SSCseverity: severity of the sickness symptom complex; SARSseverity: severe acute respiratory syndrome severity score, DIIS index: severity of critical COVID-19, SpO2: oxygen saturation; CCTA: Chest CT scan anomalies, T2DM: type 2 diabetes mellitus; BMI: body mass index

### Prediction of COVID-19 severity: results of PLS analysis

**Figure 1** depicts the results of the PLS path analysis performed on 5.000 bootstrap samples after feature selection, multi-group analysis, PLS predict analysis and prediction-oriented segmentation with critical COVID-19 illness as outcome variable. The latter was conceptualized as a latent vector exacted from ICU admission, intubation, critical illness (WHO classification) and death. With SRMR=0.028, the overall fit of this model was adequate. Furthermore, the latent vector construct reliability was adequate with Cronbach = 0.79, rho A = 0.81, composite reliability = 0.86, and AVE > 0.61 and the outer model loadings on the latent vector were all greater than 0.666, with a p-value < 0.0001. Non-significant paths were removed from this figure and the PLS analysis. We discovered that the 41.8% of the variance in critical COVID-19 could be explained by the inflammation index, CCTA, BMI, SARSseverity and age (positively associated) and SpO2 and SSCseverity (both inversely associated). CCTA, male sex (positively associated) and SpO2 (inversely associated) explained 12.2% of the variance in the inflammation index, while 23.3% of the variance in SARSseverity was explained by CCTA, age and the *NLPR3 rs10157379 CT* genotype (positively associated) and SpO2 and SSCseverity (inversely associated). We found that 19.9% of the variance in SSCseverity was explained by age and the *NLPR3 rs10754558 CG* variant (both inversely associated) and sex. There were specific indirect effects of the *NLPR3 rs10754558* variant on critical COVID-19 mediated by the path from SSCseverity to SARSseverity (t=2.32, p=0.010) and SSCseverity (t=1.98, p=0.024), yielding a significant total effect (t=2.33, p=0.010). There were specific indirect effects of the *NLPR3 rs10754558 CG* genotype on critical COVID-19 mediated by SARSseverity (t=2.16, p=0.016). There was also a significant indirect effect of the *NLPR3 rs10754558 CG* genotype on SARSseverity mediated by SSCseverity (t=2.44 p=0.007). There was a highly significant inverse effect of SSCseverity on critical COVID-19 illness (t=-5.66, p<0.0001). Male sex has significant positive effects on the outcome latent vector (t=3.73, p<0.001), which were mediated by significant specific indirect effects of sex on inflammation (t=1.94, p=0.027) and SSCseverity (t=3.54, p<0.001) and the path from SSCseverity to SARSseverity (t=3.38, p<0.001). There was a highly significant effect of age on the outcome latent vector (t=11.27, p<0.001), which was mediated by significant specific indirect effects of age on SARS (t=3.14, p=0.001), SSCseverity (t=3.07, p=0.001), and the path from SSCseverity to SARSseverity (t=4.80, p<0.001). The construct cross-validated redundancy of the critical COVID-19 latent vector (0.228) was adequate (results of blindfolding). Predicted-Oriented Segmentation analysis, in conjunction with Multi-Group Analysis and Measurement Invariance Assessment, yielded complete compositional invariance. All endogenous construct indicators had positive Q^2^ Predict values, showing that they performed better than the naivest benchmark (the prediction error was less than the error of the naivest benchmark).

**Figure 1.**
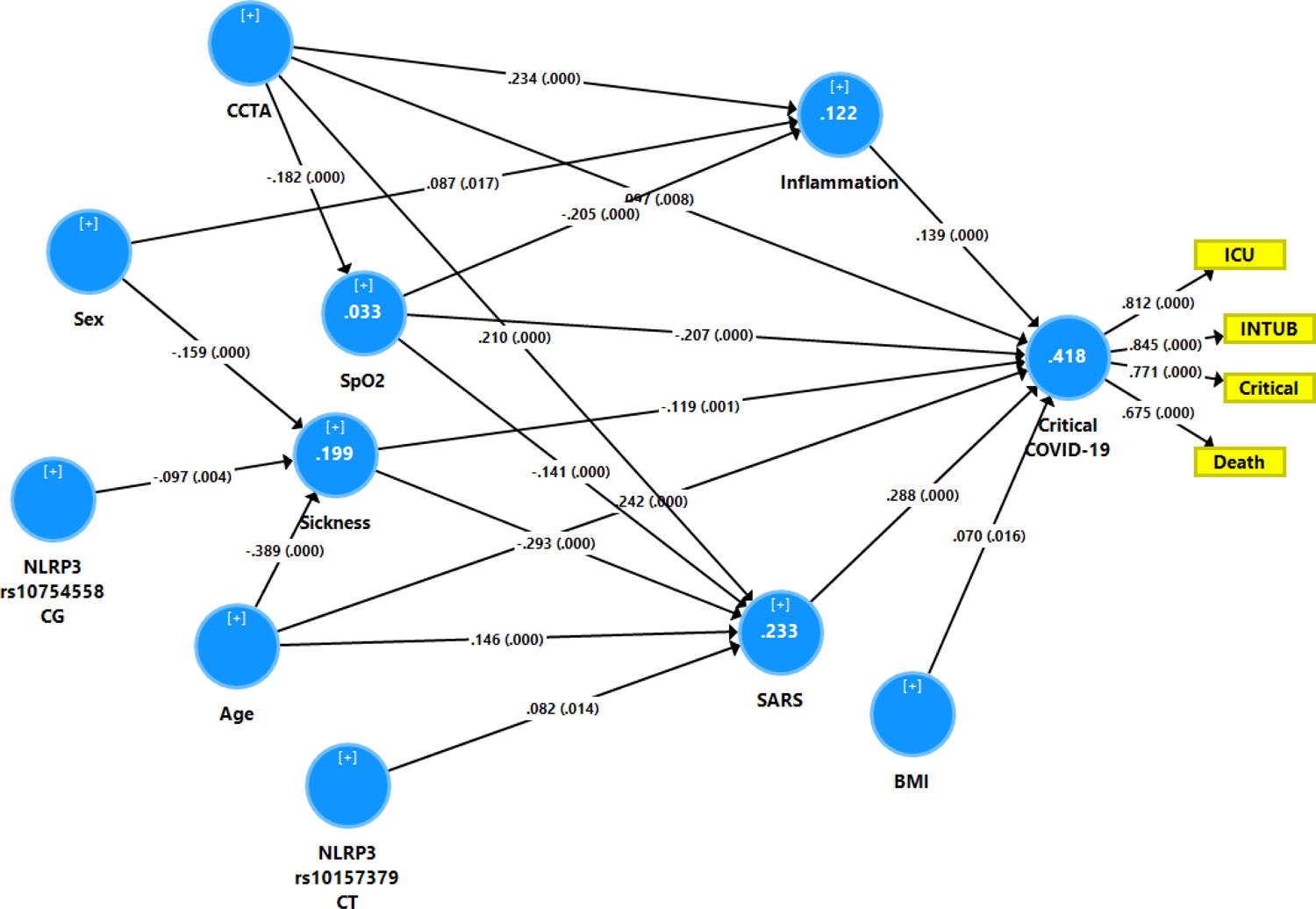
Results of Partial Least Squares analysis. ICU: intensive care unit, INTUB: intubation, critical: critical COVID-19 disease, BMI: body mass index, SARS: severe acute respiratory syndrome (SARS)-CoV-2 infection, SpO2: oxygen saturation, CCTA: chest computed tomography abnormalities, sickness: severity of the sickness symptom complex.

## Discussion

### NLRP3 SNVs and COVID-19

The first major finding of this study is that the *NLPR3 rs10157379* variant is associated with the presence of SARS, SARSseverity and severity of critical COVID-19 with the CT genotype being associated with increased SARS. Moreover, both SNVs measured in this study had cumulative effects on SARS and COVID-19 outcome whereby multiplicative effects with SSC, inflammation, CCTAs, SpO2, and BMI predicted 41.8% of the variance in COVID-19 outcome (results of PLS analysis). Our results indicate that *NLRP* SNVs participate in the response to SARS-CoV-2.

As described in the Introduction, there is now evidence that NLRP3 activation is involved in SARS-CoV-2 infection and COVID-19. Recent findings show that SARS-CoV-2 infection of monocytes [7] may prime the NLRP3 inflammasome in monocytes thereby triggering caspase-1 activation and IL-1β production, whereas selective NLRP3 inhibitors attenuate both caspase-1 activation and IL-1β levels [9]. Furthermore, the latter authors discovered that the NLRP3 inflammasome is active in the sera of COVID-19 patients, as evidenced by higher serum concentrations of Casp1p20 and IL-18, and is active in the PBMC of those patients, as evidenced by a higher percentage of FAM-YVAD+ cells, which stain active intracellular caspase-1. Importantly, the NLRP3 inflammasome is activated in postmortem lung tissues of deceased COVID-19 patients [9].

SARS-CoVs encode viroporins which interfere with cell homeostasis, contribute to the viral virulence and pathogenicity, and activate the NLRP3 inflammasome via for example lysosomal disruption leading to inflammation [20]. This causes a rapid stimulation of the innate immune response resulting in NLRP3 inflammasome pathway activation and the release of proinflammatory mediators including IL-18, IL-6, and IL-1β [21]. Furthermore, the SARS-CoV-2 N protein interacts directly with NLRP3 protein thereby promoting NLRP3 binding to ASC and facilitating NLRP3 inflammasome assembly [22]. As such, this N protein promotes the production of pro-inflammatory cytokines and aggravates acute inflammation and lung injury and, therefore, may hasten death [22]. In the acute respiratory distress syndrome, the cytokine storm in response to SARS-CoV-2 infection rather than viral replication or infection causes death in severe COVID-19 cases [21].

Most importantly, Rodrigues et al. [9] not only discovered that the NLRP3 inflammasomes is active in patients with COVID-19 but also that it may be associated with COVID-19 outcome. Thus, Casp1p20 levels but not IL-18 were increased in the severe COVID-19 phenotypes, whereas IL-18, but not Casp1p20, levels were higher in patients who required mechanical ventilation and in those with lethal COVID-19. Also, another report showed that increased IL-18 production is significantly associated with disease severity [23]. As such, the findings of the present study extent those of Rodrigues et al. [9] and Lucas et al. [23]. Nonetheless, in the present study we found no significant associations between the inflammation index, CRP or ferritin levels and the *NLRP3* SNVs, whereas Rodrigues et al. [9] reported significant associations between CRP and the NLRP3 markers Casp1p20 and/or IL-18. However, the assays of serum IL-18 and Casp1p20 are more specific biomarkers of NLRP3 activation than CRP and ferritin. All in all, the results of our study show that SNVs in the NLRP3 inflammasome may play an important role in severe and critical COVID-19 through modulating NLRP3 activity.

### Other predictors of a critical outcome of COVID-19

It should be stressed that in the current publication, critical COVID-19 was not only predicted by *NLRP3* SNVs and inflammation but also by lowered SpO2, increased CCTAs, higher age and BMI, SARSseverity, SSCseverity, and medical illness such as hypertension and T2DM. It is interesting to note that our PLS pathway analysis revealed that lung lesions and accompanying reductions in SpO2 were accompanied by increased peripheral inflammation and that cumulative effects of those three factors may worsen the outcome of COVID-19. More than 70% of individuals with COVID-19 may have CCTA abnormalities, including vascular enlargement, ground-glass opacities, bilateral abnormalities, lower lobe involvement, and posterior predilection [6]. The severity of CCTAs in COVID-19 indicates more persistent inflammation in the lungs, as well as more severe symptoms such as bronchiolitis and pneumonia, as well as lung fibrosis [24]. These lung infection sites may result in the recruitment of various immune cells, resulting in exaggerated pro-inflammatory responses [24].

SpO2 is frequently reduced in COVID-19, particularly in more severe COVID-19 cases and especially in those with increased CCTAs [6,25,26]. Lung lesions as detected with a chest-CT scan may result in decreased oxygenation [6]. Moreover, in COVID-19, lowered SpO2 is associated with immune-inflammatory biomarkers including increased levels of IL-6, IL-10, CRP, soluble receptor for advanced glycation end products, and lowered levels of albumin, magnesium and calcium [6]. Hypoxia is known to cause inflammation [27]. Therefore, our results indicate that increased CCTAs, lowered SpO2 and inflammation are intertwined phenomena which together may cause critical illness. Nevertheless, in the present study there were no significant associations between the *NLRP3* SNVs and these three interrelated factors. Our study also found that a worse COVID-19 outcome was associated with T2DM, hypertension and increased BMI. T2DM is characterized by an activated NLRP3 inflammasome as indicated by increased baseline NLRP3 activity in monocytes and increased caspase-1 activation and IL-1β and IL-18 production following stimulation of PBMCs [28, 29]. Moreover, the NLRP3 inflammasome contributes to endothelial inflammation and atherosclerosis in diabetes [30]. In Chinese patients with T2DM, the *rs10754558 NLRP3 GG* genotype is associated with insulin resistance [31]. The NLRP3 inflammasome functions as a key sensor of metabolic dysregulation, as it regulates obesity-related insulin resistance and pancreatic beta cell dysfunction explaining that NLRP3 contributes to the development of insulin resistance and T2DM [32].

A systematic review reported that the NLRP3 inflammasome is not only strongly associated with T2DM but also with obesity [33]. Interestingly, gain-of function SNPs in the *NLRP3* gene confer protection against T2DM and obesity [34]. Another recent systematic review shows that the NLRP3 inflammasome plays a crucial role in hypertension through the effects of low-grade inflammation [35]. Some *NLRP3* gene variants (e.g. *rs7512998*) are significantly associated with hypertension [36]. Consequently, we may hypothesize that the outcome of COVID-19 is aggravated by the presence of T2DM, obesity and hypertension because patients with those conditions show an activated NLRP3 inflammasome which is probably further activated by the infection. These findings extent the results of Lopes-Reyes et al. [37] who reported that SARS-CoV-2 infection may exacerbate pre-existing systemic inflammatory state of obese people by activating the NLRP3 inflammasome and releasing pro-inflammatory cytokines. These results also explain our findings that the *NLPR3* SNVs determined in our study may interact with those comorbid disorders thereby aggravating the outcome of COVID-19.

In our study, age and the *NLRP3 rs1057379* SNVs have cumulative effects on critical COVID-19 outcome and these effects were partially mediated by SARSseverity. After the age of 55 years, the risk of death due to COVID-19 increases linearly [38]. Such effects may be attributed to the effects of age on the comorbid disorders such as T2DM and hypertension and by the effects of NLRP3 inflammasome-associated inflammaging, which is accompanied by increased inflammatory mediators, AGEs, mitochondrial dysfunctions, reactive oxygen species, genomic instability, hypoxia, etc. [39].

In our study, male sex significantly and indirectly affected COVID-19 outcome by the mediating effects of SSCseverity and increased inflammation. Previously, it was shown that male sex predicts death with an OR=1.46 (95% CI 1.31-1.62) [38]. It is important to note that in male COVID-19 patients overactivation of the NLRP3 inflammasome is accompanied by increased lethality [40]. Higher plasma levels of IL-8 and IL-18 as well as stronger induction of non-classical monocytes are seen in male COVID-19 patients [41]. Testosterone may activate the NLRP3 inflammasome directly or via effects on mitochondrial ROS, while progesterone and estrogen may inhibit the inflammasome [41]. Moreover, as we will discuss in the next section, female sex strengthens SSC, which has a protective effect.

### NLRP3 SNVs, sickness behavior and COVID-19

The third major finding of this study is that the *NLRP3 rs10754558* SNV was significantly associated with SSC with the CG genotype being inversely associated with SSCseverity. Moreover, this genotype was significantly and positively associated with critical COVID-19 via the mediating effects of SSCseverity which, in turn, was inversely associated with critical COVID-19. Furthermore, SSC attenuated the presence of SARS and severe/critical COVID-19 and death due to COVID-19. These protective effects were not associated with a possible transition from the early to later phases of COVID-19 because SSCseverity was also associated with SARS and death in people with critical disease, suggesting that the protective effects of SSC are present during all stages of acute COVID-19.

Our findings indicate that SSC is in part mediated by the NLRP3 inflammasome and, thus, that the *NLRP3* SNVs may indirectly affect the outcome of COVID-19 by modulating SSC. Pro-inflammatory cytokines and especially IL-1β and IL-6 induce SSC and, therefore, the immune-inflammatory response during infection may have detrimental and protective effects [13]. Indeed, NLRP3 inflammasome activation may contribute to beneficial host-responses directed against the virus as well as detrimental effects by causing hyperinflammation [9]. The NLRP3 inflammasome may also protect against colitis, for example through the effects of IL-18 which may induce MUC2 mucin expression [42]. During Leishmania spp. infection, the NLRP3 inflammasome has not only pathogenic but also protective effects for example by driving IL-1-dependent NO-production thereby restricting parasite replication [43]. In addition, our results show that part of the protective effects of the NLRPP3 inflammasome may be mediated by SSC.

As described in the Introduction, SSC has protective effects and is part of the acute phase response and is characterized by fatigue, anergy, lethargy, myalgia, hyperesthesia, psychomotor retardation, loss of libido, anhedonia, and loss of appetite [13]. SSC is induced during the acute phase response by pro-inflammatory cytokines, including IL-6, IL-1β and TNF-α [13, 44]. For example, increased IL-1 production causes loss of appetite and other infection-associated SSC symptoms [13, 45], supporting the role of the NLRP3 inflammasome.

Moreover, headache, which is another SSC symptom, is associated with NLRP3 inflammasome activation and increased IL-6 levels [46]. SSC is a highly conserved adaptive symptom complex that protects the host by reducing the motivation to search for food so that the body can save energy to meet the increased energy requirements and hypermetabolism needed to support immune cell functions and combat acute infection [13, 47]. These symptoms keep infected animals (and humans) in a resting-like position, thereby reprogramming energy expenditure, and reducing body heat dissipation, the risk of being predated [13, 47] and social interactions and, consequently, the spread of infection through “social distancing”. During, the acute phase response, hypermetabolism increases catabolism of tissues in favor of cytokine-induced production of acute phase proteins in the liver and gluconeogenesis [13]. Most importantly, reduced foraging and nutrient intake as well as the metabolic reprogramming toward a state of negative energy balance is advantageous for the host during the acute phase response [13,47,48]. For example, in rats infected with Listeria monocytogenes, starvation prior to infection has protective effects and significantly reduces mortality [49]. Furthermore, reduced foraging may prevent elevated glucose levels, which have been linked to increased mortality in animal models of septic shock [50] and in post-stroke patients [51].

It should be noted that in the current study, we discovered that other COVID-1-associated symptoms, such as dysgeusia and anosmia are part of the SSC symptom complex. Interestingly, gastro-intestinal symptoms, diarrhea, and nausea were significantly positively associated with SSCseverity but negatively with SARSseverity. It is, therefore, possible that gastro-intestinal symptoms may contribute to decreased foraging and the sickness symptom complex by protecting against the entry of other pathogens, which may cause co-infections. This is relevant because increased levels of IL-1 and T cell activation may induce intestinal permeability, resulting in increased bacterial translocation thereby aggravating the consequences of the primary infection [52, 53], including in COVID-19 [54, 55]. As such, gastro-intestinal symptoms may be part of the SSC complex.

It is important to note that SSCseverity was not only modulated by a *NLRP3* inflammasome variant but also by age (inversely associated) and sex. Thus, SSC and SSCseverity significantly decrease with age indicating that the detrimental effects of age on COVID-19 outcome are partly related to the inhibitory effects of age on SSC. Increasing age is accompanied by overactivation of the NLRP3 explaining increased lethality due to COVID-19 [56], but not the inhibitory effects on SSC. There are, however, many other effects of age on the immune system as, for example, lowered proliferative capacity of CD4+ cells, an increase in T regulatory cells and accumulation of terminally differentiated CD8 T cells [57]. We found that female sex had a significant stimulatory effect on SSC and SSCseverity. In adults, there are many sex-linked differences in innate and adaptive immunity including increased T cell proliferation, T cell activation, TLR expression, macrophage activation, and production of pro-inflammatory cytokines in women [58]. Moreover, as discussed above, estrogens may prime the NLRP3 inflammasome. Furthermore, increased immunity to pathogens may explain why women show overall a lowered prevalence of many infectious diseases [58].

All in all, it appears that intersections among the *NLRP3 rs10754558* genetic variant, increasing age and male sex may increase risk toward critical COVID-19 by attenuating SSC. As such, lowered SSC is a new drug target especially in elderly people and males with increased BMI, hypertension, T2DM and the *NLRP3 rs10754558 CG* genotype. Future research should investigate new treatments that promote SSC via NLRP3-independent mechanisms or support nitrogen balance.

#### Limitations

It would have been more informative if we had measured NLRP3 cytokines such as IL-1β and IL-18, and caspase 1.

## Conclusions

Increased SSC is protective against SARS, and critical disease including death due to COVID-19. The *rs10754558 CG* genotype, increasing age, and male sex reduce SSC and thus lower protection against severe COVID-19. The *rs10157379 CT* and *rs10754558 GG* genotypes play an important role in SARS, and severe and critical COVID-19 especially in individuals with reduced SSC, elderly people, and those with increased BMI, hypertension, and diabetes type 2. The negative nitrogen balance that characterizes SSC is a new drug target to combat the acute phase of COVID-19.

## Data Availability

The dataset generated during and/or analyzed during the current study will be available from MM upon reasonable request and once the dataset has been fully exploited by the authors.

## Competing interests

The authors declare that they have no known competing financial interests or personal relationships that could have appeared to influence the work reported in this paper.

## Funding

There was no specific funding for this specific study.

## Author’s contributions

All authors contributed to the writing up of the paper. Statistical analyses were performed by MM. All authors revised and approved the final draft.

## Acknowledgements

None

## Compliance with Ethical Standards

### Disclosure of potential conflicts of interest

The authors have no conflicts of interest to declare that are relevant to the content of this article.

### Research involving Human Participants and/or Animals

The protocol was approved by the Institutional Research Ethics Committee of the University of Londrina, Paraná, Brazil (CAAE:31656420.0.0000.5231).

### Informed consent

Before taking part in the study, all participants and their guardians provided written informed consent.

## Notes

### Competing Interest Statement

The authors have declared no competing interest.

### Author Declarations

The protocol was approved by the Institutional Research Ethics Committee of the University of Londrina, Parana, Brazil (CAAE:31656420.0.0000.5231).

